# Meaningful outcomes of specialist leisure activities for children with complex disabilities: the views of parents, professionals and young people

**DOI:** 10.1101/2023.11.15.23298514

**Authors:** Bethan Collins, Nicole McGrath, Fiona Astill, Lisa Hurt, Sabine Maguire, Alison Kemp

## Abstract

Leisure activities during childhood are vital to quality of life and wellbeing, however parents report poor quality of life and infrequent leisure participation for children with complex disabilities. Sparkle, a charity in South Wales, delivers specialist leisure activities aimed at providing children with disabilities with access to the same opportunities as any other child. We explored the impact of this provision on psychosocial domains of quality of life for children with complex disabilities.

Multi-source qualitative case studies including interviews and observations were conducted with: four children/young people - aged 8-15 years with diagnoses including autism, Down’s syndrome and cerebral palsy - accessing Sparkle’s leisure activities; their parents/carers and leisure staff supporting them. Data were analysed using coding reliability thematic analysis.

Three themes were generated: self-development, friendship and social interaction, and self and family wellbeing. An overarching theme of the need for a specialist provision enabled the other themes linked to positive outcomes for the children.

We concluded that a specialist provision contributes to positive psychosocial outcomes linked to leisure participation for children with complex disabilities. Limitations, future research and implications for policy and practice are discussed.

## Introduction

The World Health Organisation (WHO) (2021) recently stated that the number of children with disabilities is increasing, however these children are not ‘thriving’. Understanding and improving quality of life among children and young people with disabilities is therefore an important priority. In the past few years, studies have found that children with cerebral palsy experience poorer quality of life (Makris et al., 2021), and lower life satisfaction was also found among children with additional needs, compared to those without (Rathmann et al., 2018). In order to evaluate quality of life for these children, Davis et al. (2017) identified several domains for children with cerebral palsy and intellectual disabilities, such as access to services, independence and autonomy, and social connectedness. These parameters were identified via interviews with parents; the views of the children and young people *themselves* were not included due to communication difficulties. The WHO identified six domains whilst developing their measure of quality of life: physical, psychological, level of independence, social relationships, environment, and spirituality or beliefs (WHO, 2012). Independence and psychosocial domains are common to both measures, and this paper focuses on these domains of quality of life, including positive feelings, self-esteem, and social relationships, which contribute to psychosocial wellbeing.

Leisure activities during childhood are vital to the development of social relationships, and are important opportunities for skills development and to experience positive emotions, all of which significantly impact the psychosocial wellbeing of children and young people (Brajsa-Zganec et al., 2011; Merrells et al., 2018). National Institute for Health and Care Excellence guidance reminds us that *“social participation is as important as care and education for maintaining and improving the quality of life of disabled children and young people with severe complex needs”* (NICE, 2022). Indeed, Shikako-Thomas et al. (2012) found frequent participation in physical leisure activities was related to improved physical and psychological wellbeing for children with cerebral palsy. Longo et al.’s (2017) findings support this: frequent and diverse participation in leisure activities led to improvements in physical wellbeing and social acceptance and support, and enjoyment of leisure activities was related to positive feelings and life satisfaction.

Frequent participation in leisure activities is related to better overall quality of life, however parents report *poor* quality of life and *infrequent* leisure participation for children with disabilities who have the most complex mobility, communication and personal care needs (Williams et al., 2021). These children often experience barriers to accessing leisure opportunities, including judgement and bullying (Brooks et al., 2021; Merrells et al., 2018). Those with the most complex needs, such as severe communication difficulties, seizures, and those who rely on others for personal care, are most likely to be excluded (Bult et al., 2011; Merrells et al., 2018).

Sparkle is a charity supporting children and young people, aged 0-17, with complex disabilities in Gwent, South Wales, UK. As the charity partner to three health board-run disabled children’s centres – Serennu, Nevill Hall and Caerphilly – Sparkle aims to enhance statutory health and social care services with family and child support. Sparkle delivers specialist leisure activities from the centres and other appropriate venues in their catchment areas, including weekly play and youth clubs. The aim of this provision is to provide children and young people with disabilities with the same leisure opportunities as other children, and support to develop social skills and independence (Appendix 1). The provision also provides short respite opportunities for families and caregivers. More detailed information about the provision can be found in figures 1 and 2.

**Figure 1:**
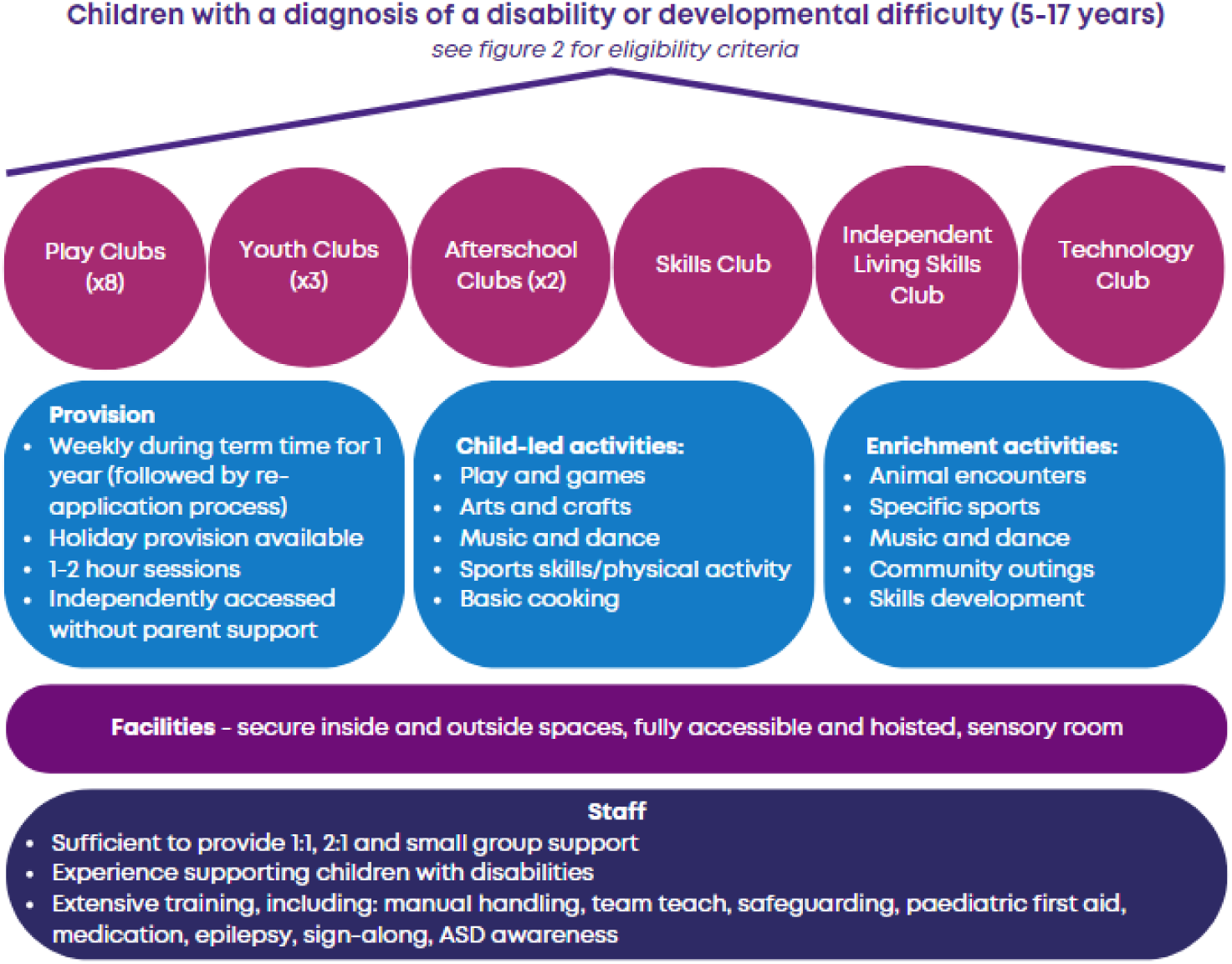
Specialist leisure provision for children aged 5-17 years with complex disabilities, provided by Sparkle (South Wales).

**Figure 2:**
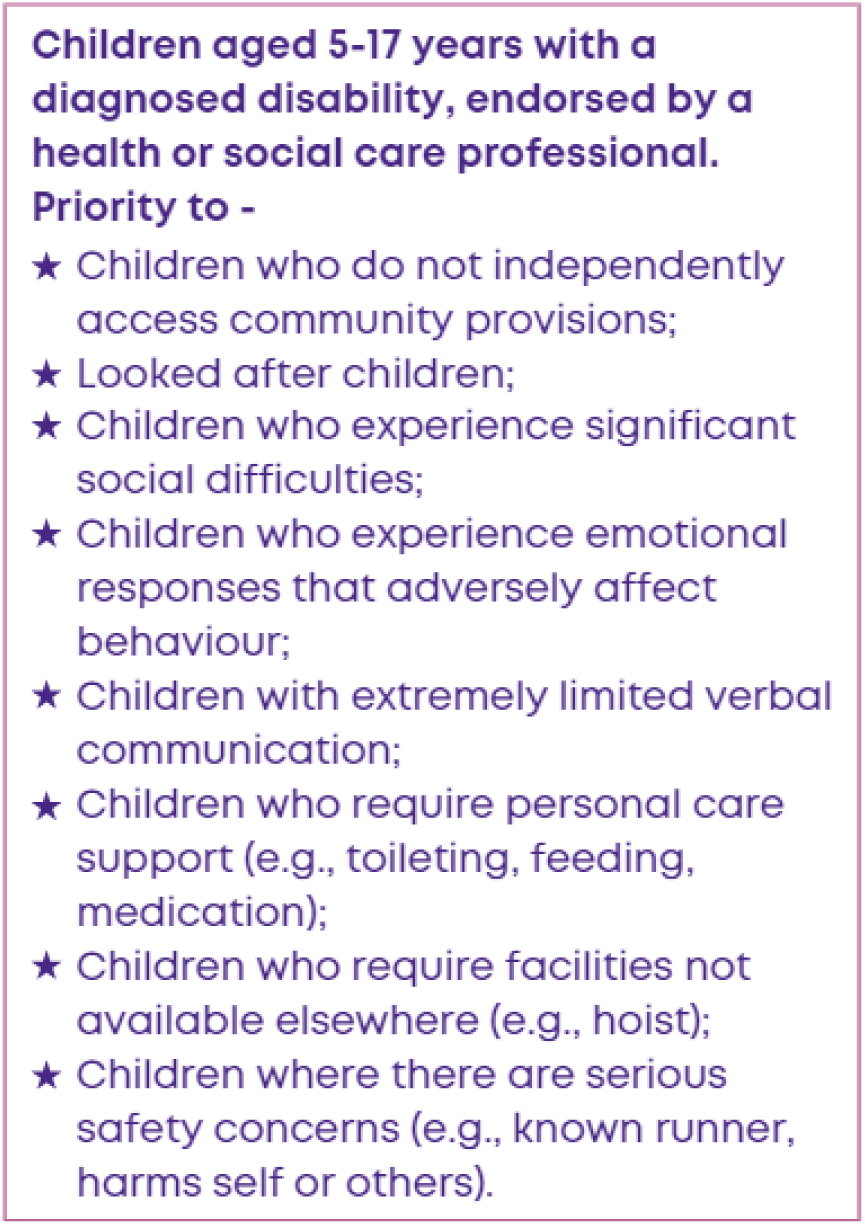
Eligibility criteria for Sparkle (South Wales)’s specialist leisure provision for children aged 5-17 years with complex disabilities.

NICE (2022) recommends research into the components of ‘short break’, or respite, services that are effective for children and young people with complex disabilities, and their families. Sparkle has previously aimed to evaluate the impact of this specialist leisure provision using quantitative tools aimed at measuring quality of life in children with disabilities. One parent-report measure found a significant change in the ‘positive emotions’ domain of quality of life for children after accessing the leisure provision for 6 months, however there was no significant change in the other domains or overall quality of life (McGrath et al., 2023). Tools aimed at including the children and young people’s views were not found to be suitable for the diverse population served by the provision (Astill et al., 2023). Makris et al. (2021) suggests that self-report should be relied on due to the subjective nature of quality of life, and the authors propose that a parent/carer’s own distress and anxieties surrounding their child’s disability could influence proxy-report scores. Including the views of children and young people, parents/carers and professionals to provide information from various contexts was recommended by Makris et al. (2021) as a ‘gold standard’ approach.

We have therefore conducted a pilot multi-source case study model to evaluate and explore the impact of Sparkle’s specialist leisure provision on psychosocial domains of quality of life for children and young people with complex disabilities. We anticipate that this study will inform practice, policy and future research direction and methodology.

## Materials and Methods

### Design

In depth qualitative case studies with four children/young people and their families were conducted, using three separate sources of information.

### Participants

Participants for the case studies were recruited using purposive sampling from families accessing Sparkle’s specialist leisure activities. Parents/carers were invited via email, and children of parents/carers who responded to the email were invited verbally during the provision. Inclusion criteria were children and young people between 8 and 16 years old who had accessed Sparkle leisure activities for at least one year. This age range was chosen to include children old enough to engage in the evaluation, and to exclude those older than 16 completed years who were nearing transition out of the service. We aimed to include both male and female children and young people, with various diagnoses and communication styles, attending different leisure clubs; 16 families were invited and four participated. Few invited parents/carers gave a reason for not wanting to participate, however one reason given was that they did not believe their child would be able to answer any questions or take part in the evaluation (this reason came from parents/carers of children with limited or no verbal communication skills). Data collection ceased when a representative sample was achieved.

### Data collection

Four in-depth case studies were conducted using qualitative methods, including semi-structured interviews with a *parent/carer*, interviews with *Sparkle leisure staff* supporting the children, and observations of the *children/young people* during the provision, complemented by informal conversations with children/young people tailored to their communication style (see figure 3). Interview schedules for parents/carers and leisure staff, and an observation schedule, were formulated by the researchers (Appendix 2). Data were collected between January and May 2022. Interviews with parents/carers were conducted via Microsoft Teams or telephone, by either Sparkle’s Research and Development Officer, who has a master’s degree in psychology and experience of conducting qualitative research for charities, or an Aneurin Bevan University Health Board Paediatric Registrar, who has research experience and no formal affiliation with Sparkle. Interviews were audio recorded with participants’ permission and lasted on average 30 minutes. Interviews with leisure staff supporting the children, and observations of the children during leisure activities, were conducted at Serennu Children’s Centre by Sparkle’s Research and Development Officer. Interviews with leisure staff lasted on average 10 minutes. One leisure session, lasting 1 or 2 hours, was observed for each child/young person; the observer made hand-written notes following the observation schedule and wrote down comments made by the children/young people.

**Figure 3:**
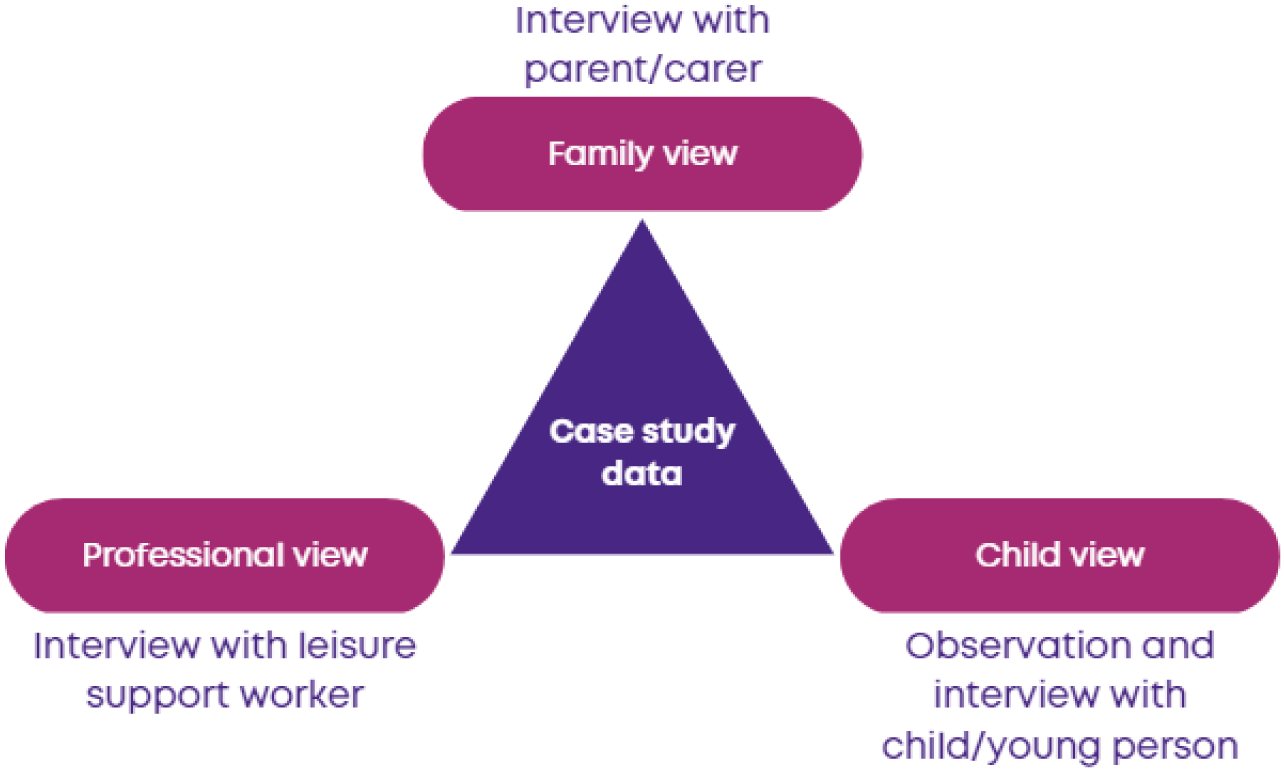
Data collection sources for in-depth case studies of children with complex disabilities accessing a specialist leisure provision.

### Analysis

Interview transcripts and observation notes from the case studies were analysed using qualitative data analysis software, NVivo (QSR International). Coding reliability thematic analysis was used to provide topic summaries; transcripts were coded inductively to identify, analyse and report patterns with the data (Braun and Clarke, 2022). The data was coded by Sparkle’s Research and Development Officer, and codes were independently reviewed and agreed by two Aneurin Bevan University Health Board Paediatric Specialist Registrars who have no affiliation with Sparkle.

### Ethical approval

This service evaluation was approved by Aneurin Bevan University Health Board Research and Development Department Research Risk Review Panel on 6^th^ December 2021. Informed written or verbal consent was obtained from all participants.

## Results

Themes generated from four in-depth, qualitative case studies are discussed below. Two boys and two girls, aged between 8 and 15 years, participated; diagnoses included autism, Down’s syndrome and cerebral palsy. Three children communicated verbally whilst one was nonverbal. Two of the children required 1:1 support from a Leisure Support Worker, whilst the other two were supported as part of small groups within their clubs (1 staff member supporting up to 4 children). Themes were generated by combining data from the interviews with parents/carers, leisure staff and the children and young people, as well as researcher observations. We identified whether quotes came from parents, Sparkle staff, researcher observations or comments from the children or young people, as well as whether the quote relates to child/young person A, B, C or D.

### Self-development

Participation in clubs contributed to multiple areas of skills development and social relationships among children and young people. This was highlighted in the first theme. The specialist provision enhanced the children and young people’s development, as staff understood that the young people needed encouragement and patient support to reach targets.

> *“Often took part in activities if prompted or encouraged and supported by staff. Withdraws from activities she does not enjoy.”* (Researcher observation, Child A)

This understanding and support allowed young people to achieve significant milestones.

> *“Child D is non-verbal but was making noises and said a few short words during the session, which staff said was new and seemed to be a significant milestone.”* (Researcher observation, Child D)

Leisure staff reported achievements against goals set for the children, including improved communication skills, increased interaction and participation, and being able to self-regulate their emotions.

> *“Communication has come on a lot and she’s more likely to say what she wants, whereas before she would refuse to answer or choose when asked what she wants.”* (Sparkle staff, Child C).
>
> *“He tries to calm himself down now, it’s just the frustration that builds up in him really. He does try to socialise with others now, as well as the few friends he has.”* (Sparkle staff, Child B)

The young people gained confidence from accessing the provision.

> *“She’s not afraid to talk to anyone or play, I think she’s really comfortable in club. She loves all the staff, she has one or two close friends in the club but isn’t afraid to join in with a group. […] She’s really confident, not afraid to talk to you, loves talking about her day.”* (Sparkle staff, Child A)

Parents commented that their child accessing the provision encouraged their independence, by being able to access leisure activities without needing parental support.

> *“We’ve always done a lot of groups and things, when she was younger we used to do like gymnastics and things, like toddler things, and up to a certain age that’s fine. But now, the difficulty with that is that out of school, all activities and anything like that, that’s not always realistic without that one-to-one support […] So I think having that independence to go to groups and go to different things, without necessarily needing me.”* (Parent, Child A)
>
> *“Life skills. The whole aim is, it was to try and get her to interact and build friendships and get out of the house, doing something and being active rather than just being around us, because that’s all she wanted to do was just be around me all the time.”* (Parent, Child C)

The provision encouraged self-agency for the children; they were encouraged to choose or plan their own activities and explore their own interests, promoting their independence and self-efficacy.

> *“I choose.”* (Young person during researcher observation, Child A)
>
> *“Last week we surveyed what they would like to cook and they chose these mint choc chip cupcakes so that’s in place for them to do this week. […] But it’s them voicing what they want to do, like if someone wants to make a card for something we’ll go down the craft cupboard and adapt.”* (Sparkle staff, Child B)
>
> *“Child C went with a member of staff and another young person to collect a variety of games for the group to choose from.”* (Researcher observation, Child C)
>
> *“We’ll set out toys in all the different rooms and the children are just allowed to like wander and choose, we’re not telling them to do anything so it’s completely up to them what they want to do.”* (Sparkle staff, Child D)

### Friendship and social interaction

Parents thought that friendships developed at this specialist provision were not forced, whereas in mainstream settings such as school, they felt social interaction was orchestrated by teachers or typically young people were ‘looking after’ their children, rather than developing a friendship on ‘equal footing’.

> *“The main thing is because she goes to mainstream school – as much as school are very keen, they say like everybody loves her, they all take turns to play with her, she has friends - I think the reality is that as she gets older, they’re not necessarily meaningful friendships […] I wanted her to mix with other children with additional needs to make those friendships on more of an equal footing I suppose.”* (Parent, Child A)
>
> *“In his primary school he didn’t really have any friends. A couple of the girls were friends with him but they used to mother him. He had trouble bonding with any of the boys because he didn’t really like football, he couldn’t really do football because he couldn’t run around fast as what they could.”* (Parent, Child B)

The development of meaningful friendships was indicated by children and young people who were able to differentiate between those in club they considered friends and the other children.

> *“Do you like playing with others or by yourself?” “With friends” “Who are you friends?” Child A lists friends by name, and can differentiate between her friends and other children in club. “What is your favourite thing about club?” “Friends”* (Young person during researcher observation, Child A)
>
> *“She was telling my sister yesterday morning that she was going to club, she was going to play with her new friends and she knows the names of her friends.”* (Parent, Child A)
>
> *“Child C names one friend, and differentiates between her friend and other children in club.* (Researcher observation, Child C)

Participants seemed to agree that the children being with their peers, i.e., other children with disabilities, facilitated social development at their own pace. For some children this meant learning to tolerate other children playing nearby, whilst for others it meant developing meaningful friendships and a ‘social life’.

> *“[He] wouldn’t have had a social life if it wasn’t for [club].”* (Parent, Child B)
>
> *“Child A introduced herself to me without prompting and straight away eagerly interacts with staff. Will interact with some other children, however seems to require support from Leisure Support Worker during interactions.”* (Researcher observation, Child A)

Young people who struggled to interact with the other children gained social skills by interacting with staff.

> *“She has a good relationship with one member of staff and one young person in the group and can be chatty with them.”* (Sparkle staff, Child C)
>
> *“I think he is interacting with others because the staff can understand him better, he doesn’t make any friends, even at the school he doesn’t. Most of the time the interaction is with the [Sparkle staff].”* (Parent, Child D)

### Self and family wellbeing

Positive responses regarding accessing the clubs were shared by both children and young people and their families. Accessing the clubs was noted to impact wider family wellbeing, alongside the wellbeing of the children and young people themselves. Observations and comments from the children and young people demonstrated that they enjoyed the provision and wanted to be there.

> *“Child B showed signs of enjoyment, including smiling and laughing.”* (Researcher observation, Child B)
>
> *“How do you feel at club?” “Nice”* (Young person during researcher observation, Child C)
>
> *“Showed signs of enjoyment, such as smiling and laughing, when running up and down the corridor playing a chase game with Sparkle staff.”* (Researcher observation, Child D)

Parents described the children and young people as being happier since accessing the provision.

> *“As long as she’s enjoying herself that’s all I want her to do, she wants to go and that’s the biggest part of it, that she wants to go. […] She comes home and she’s raving, she’s like we did this and we did that. […] She just loves it.”* (Parent, Child C)
>
> *“He is quite relaxed after going there, he’s quite happy when we go there, it’s an hour and he is quite happy. In the evening, he is relaxing after it, but if we don’t take him to club, like if we went on a trip, he’ll miss it.”* (Parent, Child D)

This improved mood had an impact on the family’s wellbeing; parents were happy that their child was happy, and described improved home environments when their child was in a good mood and wanted to tell them about what they did during the session.

> *“It’s a nicer relationship, she comes home happy and wants to tell me all about it, the mood is completely different. […] the change in her has impacted on all of us, there’s less arguments, the mood is different, happier.”* (Parent, Child C)

Families said that their child was calmer following sessions, allowing them to spend quality time together at home.

> *“After he goes and plays, he is calm for one or two hours, and that is quite beneficial for us and him and at least he is safe for 1 hour and I can do something, focus on something else, so it’s good for myself as well. […] when he is calm we have the chance to be together and talk to each other and enjoy our time.”* (Parent, Child D)

Carers and siblings’ wellbeing benefitted from respite time whilst the children and young people were accessing the provision, allowing them to ‘take a break’.

> *“Actually, it is really a relief to have somewhere that we know we can take him, even to leave like a normal child it is really hard to find a childminder to look after, with this sort of children to leave him for even 1 hour to go shopping or somewhere, because I need to be with him all the time.”* (Parent, Child D)
>
> *“This was the first time I sent her during the Christmas holidays, but I wouldn’t realistically send her to a play scheme in school or a mainstream place I wouldn’t send her to, whereas those couple of hours in the holidays were a couple of hours I could do something with her sister.”* (Parent, Child A)

### Need for specialist provision

Throughout the case studies, a prominent theme of the service offering a safe space and the children requiring a specialist provision was evident (see figure 4).

**Figure 4:**
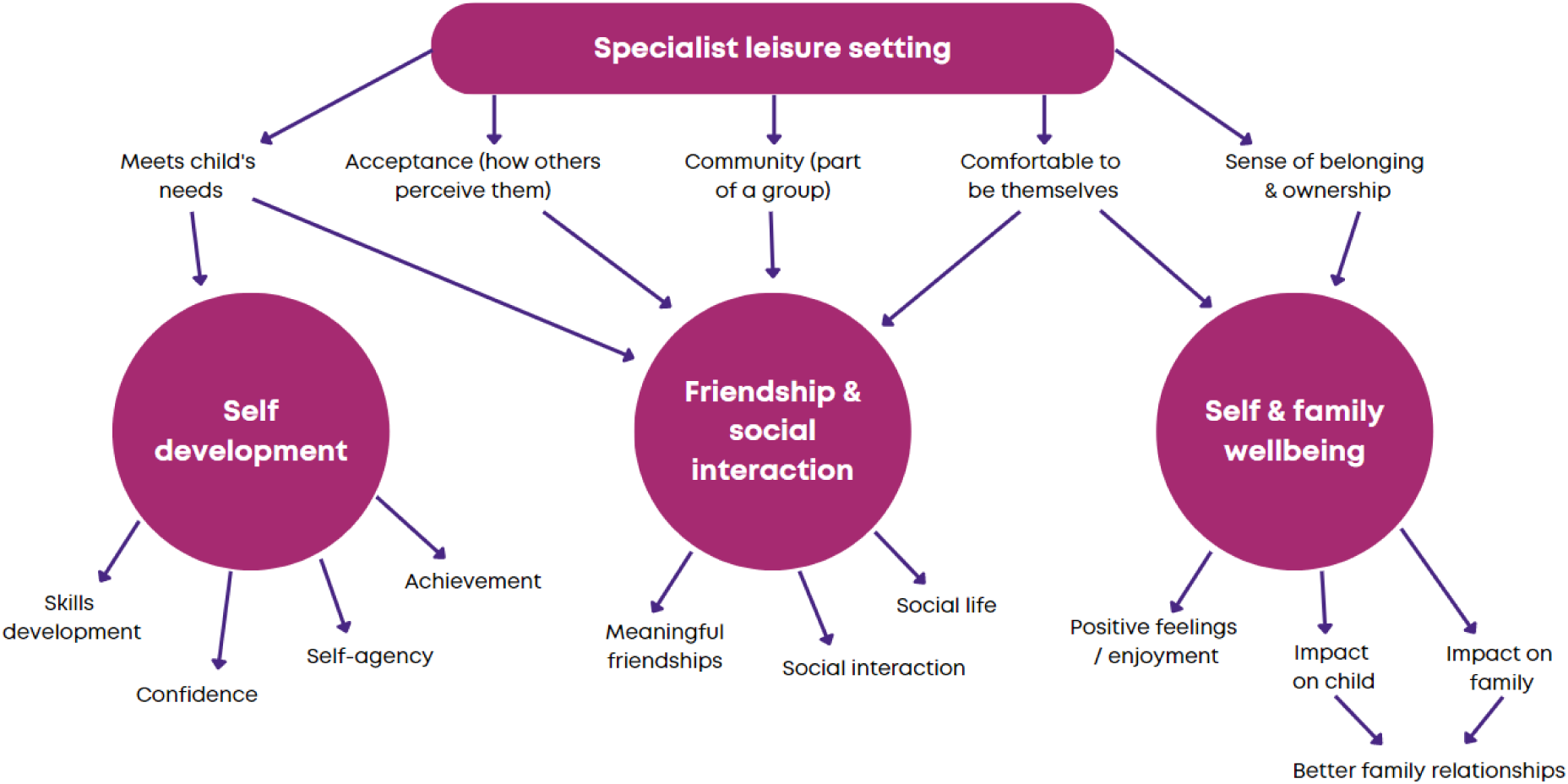
Codes and themes derived from in-depth case studies of children with complex disabilities accessing a specialist leisure provision.

> *“He needs 100% attention, there must be someone with him all the time, otherwise what will happen is he will get quite distressed and try to do destructive things and his behaviour is quite challenging. […] He also doesn’t have any awareness about danger. […] He gets really distressed most of the time because he can’t express himself, because he can’t speak.”* (Parent, Child D)
>
> *“We just wanted her to have something, she’s joined clubs before but because of her processing it ends up, she doesn’t like it because she gets left behind, they don’t understand that she needs someone to explain what’s going on, explain the rules of the game, encourage her to talk to someone or get involved.”* (Parent, Child C)

Due to their disabilities, the children and young people supported by this specialist leisure provision have complex behavioural, processing and communication needs. Parents commented that their children needed a secure setting with specially trained staff for them to be able to access leisure provision.

> *“It just gives you that completely different mind-set, you know that she’s with people that know how to deal with situations that come up […] with the best will in the world if you’re not experienced with children with additional needs, you don’t necessarily know how to deal with that situation. It makes a big difference to think that she can go there and you know that she’s safe and she’s happy.”* (Parent, Child A)
>
> *“I think it’s definitely working keeping the same staff member with him […] I’d say just how he is now he would always need a one-to-one to support him and he doesn’t want to interact with other children, right now they scare him. So I couldn’t imagine him being able to manage in a mainstream setting.”* (Sparkle staff, Child D)

The service offered a space where everyone understood the children’s needs, so families didn’t feel that they were being ‘judged’.

> *“We can’t go to a normal park or soft play and café or anywhere, he doesn’t know how to behave according to those, you know the covid rules and all, and the people don’t know why he’s behaving, the normal people who are coming there are distressed because of him and we are distressed because we couldn’t make him understand.”* (Parent, Child D)

This resulted in Sparkle clubs offering a community where the children and young people could feel comfortable to be themselves, whereas they might not have felt this way in mainstream settings.

> *“It makes things a lot easier, because you haven’t got to try and integrate him into other situations where there’s children the same age but they haven’t got a diagnosis. You’re not, how can I put it; you’re not apologising for him to be completely honest. Sometimes where he is a bit loud and a bit out there around other kids and they could think he’s naughty, but he’s not naughty he is just him. So yeah you’re not apologising for things in Sparkle, and we’re all in the same boat. So it’s hard being with kids without disabilities, but the ones that he’s with in the club, everyone is the same so it’s like a community, so that’s the good thing about Sparkle.”* (Parent, Child B)
>
> *“It’s how other children would perceive him. I could see him being possibly an easy target for some mainstream children […] He’s very imaginative in his play and they would possibly ridicule him for one of his favourite activities and then he wouldn’t feel comfortable doing that.”* (Play Worker, Child B)

This facilitated a sense of belonging and ownership regarding the service, as the young people had a place and a provision that was only theirs.

> *“It’s really helped him build his confidence and just to interact with others, because he doesn’t have any chances to interact with others, because of his behaviour and he is not verbal, it is quite difficult for him to be with others, but the club gives him kind of a place for him to be, other than the school and the family.”* (Parent, Child D)
>
> *“This is my home, mine and [my friend]’s home.”* (Young person, Child B)

### Consistency between data sources

The consistency between data sources for each child/young person throughout the themes supports the validity of these findings. Of particular note, findings around the development of meaningful friendships were echoed by parents/carers, leisure staff and the young people – parents/carers discussed wanting their child to develop meaningful friendships ‘on an equal footing’, whilst children and young people could differentiate between those they considered friends and other children attending the provision, and leisure staff also commented on the friendships young people were developing. Similarly, both parents/carers and leisure staff commented on the provision supporting the children to participate in more activities via encouragement and tailoring activities to meet processing needs, which was also captured by the observations. A sense of community and belonging was brought up by parents/carers, leisure staff and young people individually; parents/carers felt their children were not judged and could be themselves at the specialist provision, which leisure staff also commented on and shared their fears that the same could not be said if they accessed a mainstream provision, and the young people described the provision as their ‘home’.

## Discussion

This multi-source case study evaluation shows that specialist leisure provisions have the potential to improve the psychosocial wellbeing of children with complex disabilities. Improvement in self-development, friendships and social interaction, and self and family wellbeing were all described by the children, their parent or carer and leisure staff at Sparkle. The benefits of the specialist provision were constantly highlighted by parents, together with the risks and challenges posed by inappropriate community provision.

The activities undertaken within and between the clubs varied. However, the ethos of the clubs was consistent. Clubs were held regularly (weekly), the leisure activities were child-led within a group of children with complex disabilities, in a parent-free environment. The leisure facilities were secure and run by experienced, well-trained staff, with a low staff-to-child ratio. This specialist leisure setting enabled the three themes (self-development, friendship and social interaction, and self and family wellbeing) linked to positive outcomes for the children and their families.

Within each of the three themes significant benefits were characterised. The children and young people showed greater levels of confidence, achievement and independence. They were able to exhibit their own choices of activities and interact and participate with their peers within the clubs. Friendships developed were described as meaningful. The children were described as happier and calmer, moods that often extended beyond club into family life. Parents and carers had the reassurance that children were in a safe environment, where they were comfortable and experienced a sense of belonging.

Research into the differing impacts of specialist versus integrated leisure is limited. Some studies suggest that there is no difference in self-concept among teens and adults with intellectual disabilities accessing mainstream and specialist leisure activities (Merrells et al., 2018; Duvdevany, 2002). However, young people with disabilities want to access leisure somewhere where they feel safe and unjudged (Brooks et al., 2021). A study by Merrells et al. (2019) which involved qualitative interviews with young adults, aged 18-24, with disabilities described participants being excluded from community leisure and social opportunities and being seen as ‘different’ – however, leisure provisions designated specifically for people with disabilities resulted in enjoyment and a feeling of belonging to part of a community. These specialist leisure activities provided opportunities to form friendships or spend time with others that they relate to, as well as providing them with a safe space, which echoes the findings of the current evaluation of a specialist leisure provision for children with disabilities.

A key finding from this evaluation was that a specialist leisure provision for children and young people with complex disabilities delivers meaningful and positive outcomes. This is a particularly important finding given current debates surrounding specialist versus inclusive placements for children with disabilities. This discussion is usually related to education placements, and the United Nations Convention on the Rights of Persons with Disabilities champions inclusive education for children with disabilities (Vyrastekova, 2021; Kramer et al., 2021). Buchner and Thompson (2021) discussed how there has been a shift from specialist, segregated education for children with disabilities to inclusive education being regarded as best practice globally. However, they highlight problems such as a ‘lack of strategies to reduce numbers in special schools’, and whilst some countries introduced ‘one curriculum for all’ schemes, these have been challenged and separate curriculums for children with intellectual disabilities developed. Despite this, Wehmeyer et al. (2021) suggests inclusive placements should be the default for children with disabilities, due to potential positive benefits such as improved academic achievement, communication skills, social interaction and perceptions of belonging. Kramer et al. (2021) found students with mild learning difficulties in inclusive settings experienced better academic and cognitive outcomes compared to those attending specialist schools, however there was no difference in psychosocial outcomes between the two settings, and no mention of children with more complex disabilities.

Research into the effects of specialist versus inclusive education on social and personal outcomes is mixed; whilst Vyrastekova (2021) and Hardiman et al. (2009) found no difference between specialist and inclusive schools in how lonely parents perceived their child to be or children’s social competence, one study found life satisfaction among those with additional needs was higher at special schools than mainstream schools (Rathmann et al., 2018). A meta-analysis by Dalgaard et al. (2021) found inconsistent effects of inclusive education on academic and psychosocial outcomes for children with additional needs, with no evidence to suggest placing children with additional needs in inclusive schools by default was superior to specialist schools. It was recommended that individual assessments of each child’s needs are required, rather than a ‘one size fits all’ approach.

This evaluation was limited by a lack of comparison with children and young people with disabilities accessing mainstream leisure provision, or not accessing any leisure provision. However, some parent participants did comment on previous attempts for their child to access a mainstream provision, their inability to access due to inadequate provisions of the necessary facilities or support to meet complex needs, and the difference between these attempts and the specialist provision made by Sparkle. The analysis of data from the four participants may have been biased towards their perceived benefits of the leisure facilities versus the 12 families who did not participate in the study. There was the potential for bias in the evaluation, as one of the researchers collecting and analysing the data was an employee of Sparkle; however, this was counteracted by independent researchers reviewing and agreeing the thematic coding during analysis, and the consistency of findings between the various sources of data. The leisure provision evaluated is a real-world, active service, and evaluations of such services present a variety of challenges, such as around participant recruitment and lack of control groups, and there are particular challenges associated with including this population in research due to complex communication and cognitive difficulties. However, despite these challenges, it is important that real-world services are evaluated to ensure they meet the needs of the children and young people, and that their views are heard.

A number of the studies cited in this report did not include the direct views of children and young people due to their complex needs, cognitive or communication skills; some researchers excluded children with the most complex disabilities completely from their studies (Davis et al., 2017; Shikako-Thomas et al., 2012; Longo et al., 2017; Kramer et al., 2021; Dalgaard et al., 2022). Whilst the current evaluation used a method involving input from multiple sources, as recommended by Makris et al. (2021) as a gold standard approach, input from the children and young people was limited, particularly from those with severe communication difficulties. However, the consistency between the parent/carer, child and leisure staff comments, and observations of the individual children and young people, reinforced our confidence in the methodology and the findings.

There is a clear need for further research into the impact of specialist leisure provision for children and young people with complex disabilities, compared to inclusive or mainstream provisions, not least as it is a more expensive provision. Further research in this area could have significant implications for policy and service delivery. This multisource case study method can be developed to increase opportunities for children and young people with complex disabilities to share their views. Future evaluations of services for children with disabilities could include measures of self-development, friendship and social interaction, self and family wellbeing, and the leisure environment meeting the specialist needs of the child, which this research suggests contribute to psychosocial wellbeing.

### Conclusions

This multisource evaluation of a leisure provision for children and young people with complex disabilities found that a specialist setting results in positive psychosocial outcomes for the children, linked to self-development, friendships and social interactions, and self and family wellbeing. The findings highlight an important issue relevant to current debates on specialist versus inclusive provisions for children with disabilities, and the research includes the direct views of children who are often excluded from studies due to their extremely complex communication and behavioural needs.

## Acknowledgments

The authors would like to thank the children, young people, parents and Sparkle leisure staff who supported and participated in this evaluation.

## Data availability

Transcripts and observation notes are not openly available due to concerns about potential identifiability and lack of participant consent to open the data.

## About the authors

Bethan Collins MSc is the Research and Development Officer at Sparkle (South Wales), based at Serennu Children’s Centre in Newport, South Wales. Dr Nicole McGrath is a Consultant Paediatrician at Aneurin Bevan University Health Board, based at Serennu Children’s Centre. Dr Fiona Astill is a ST6 Community Child Health Registrar at Aneurin Bevan University Health Board, based at St Cadoc’s Hospital. Dr Lisa Hurt is a Senior Lecturer in maternal and child health epidemiology at Cardiff University School of Medicine, Division of Population Medicine. Dr Sabine Maguire is an Honorary Research Fellow at Cardiff University, and an unsalaried trustee of Sparkle (South Wales). Professor Alison Kemp is an Emeritus Professor at Cardiff University School of Medicine.

## Appendix 1 Aims of the specialist leisure clubs delivered by Sparkle (South Wales).

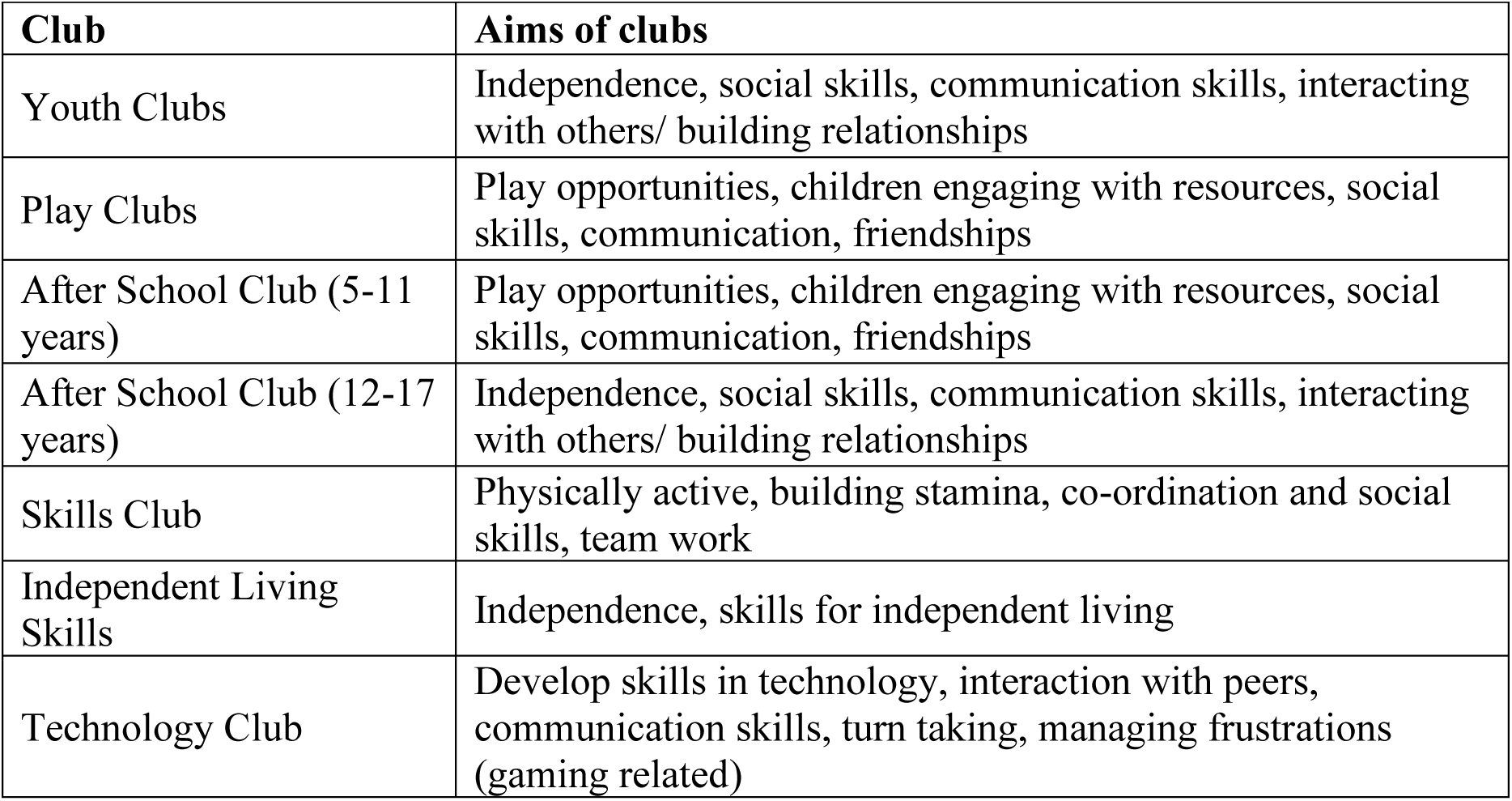

## Appendix 2 Interview and Observation Schedules for Case Studies Evaluating Specialist Leisure Provision for Children with Disabilities

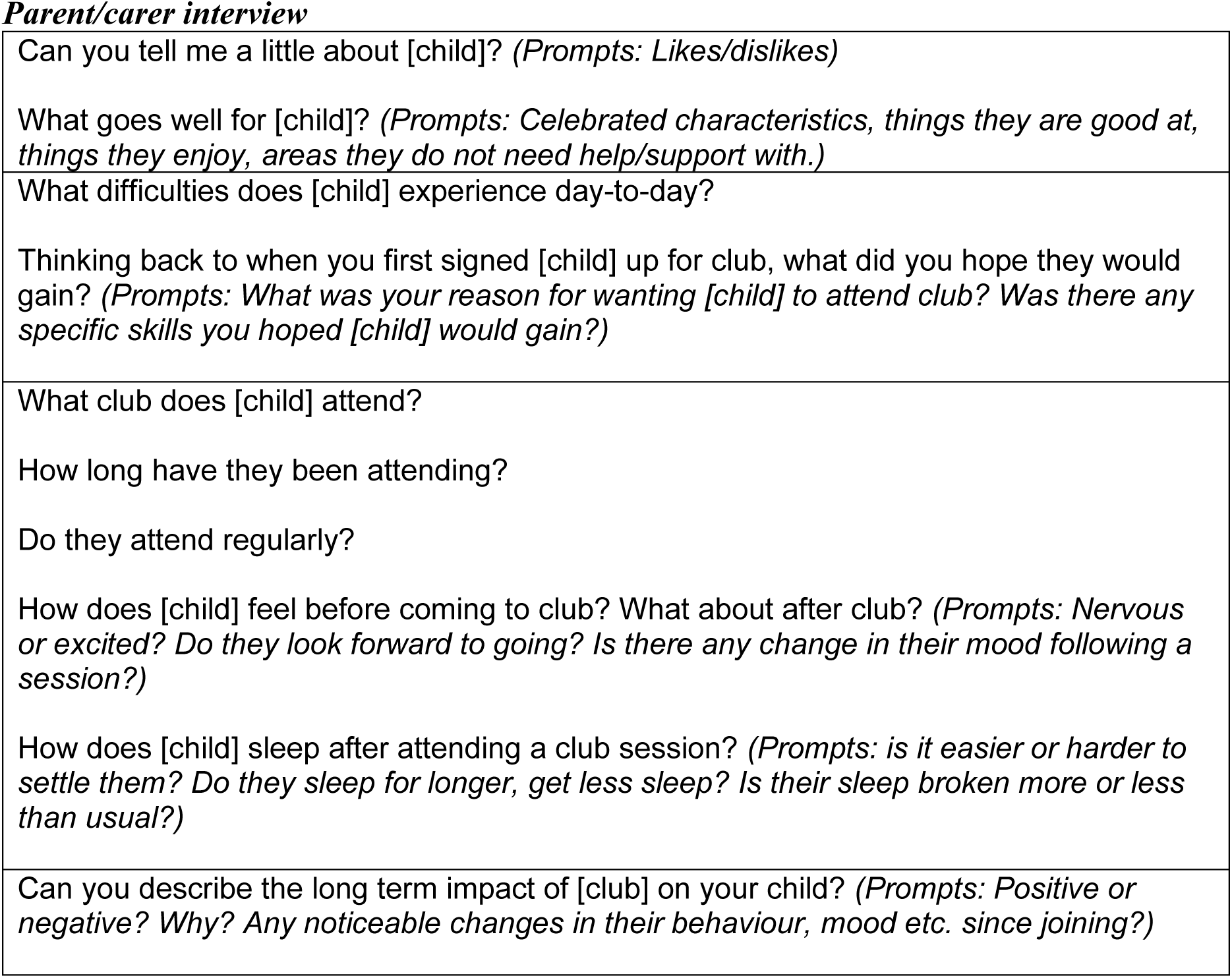
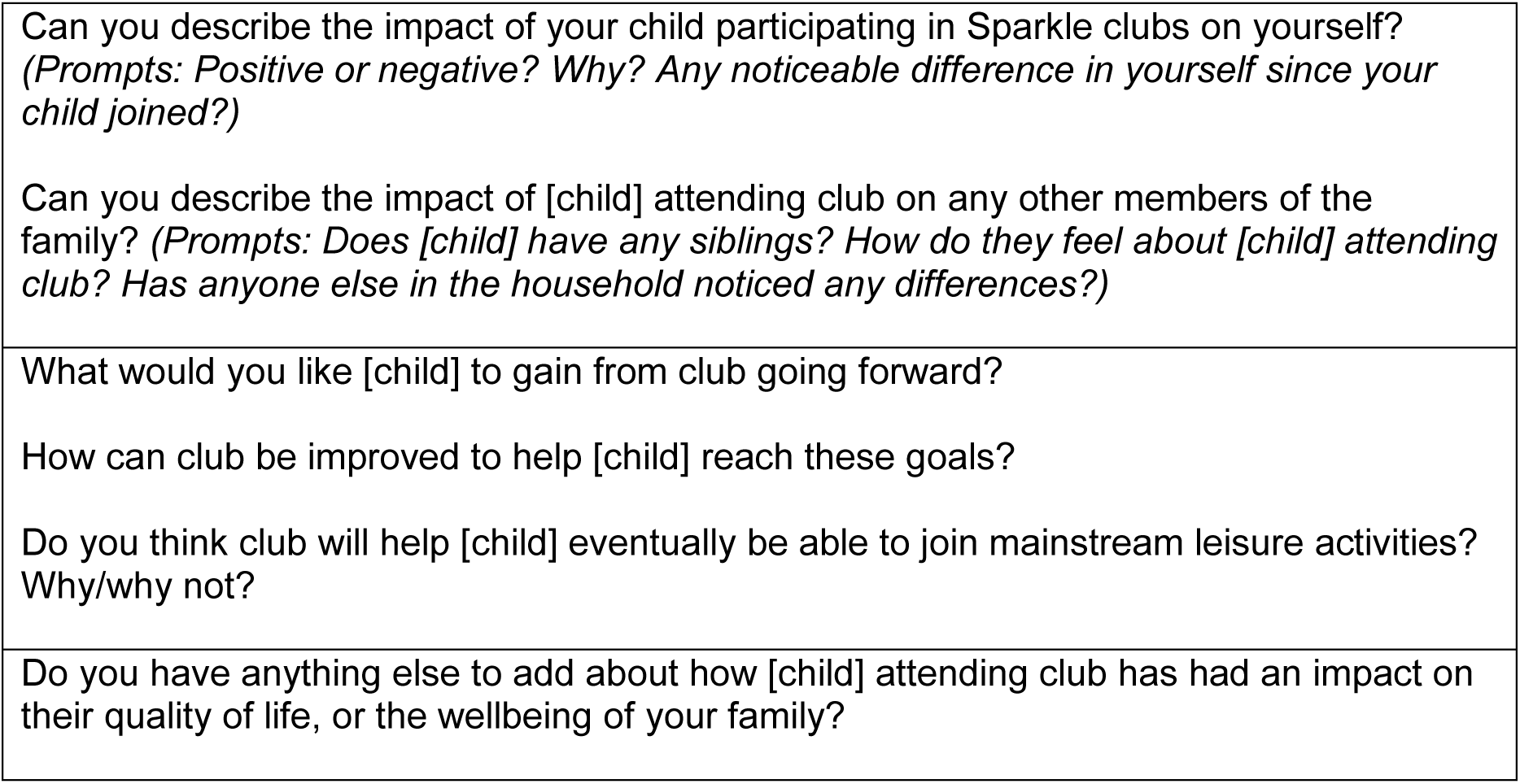
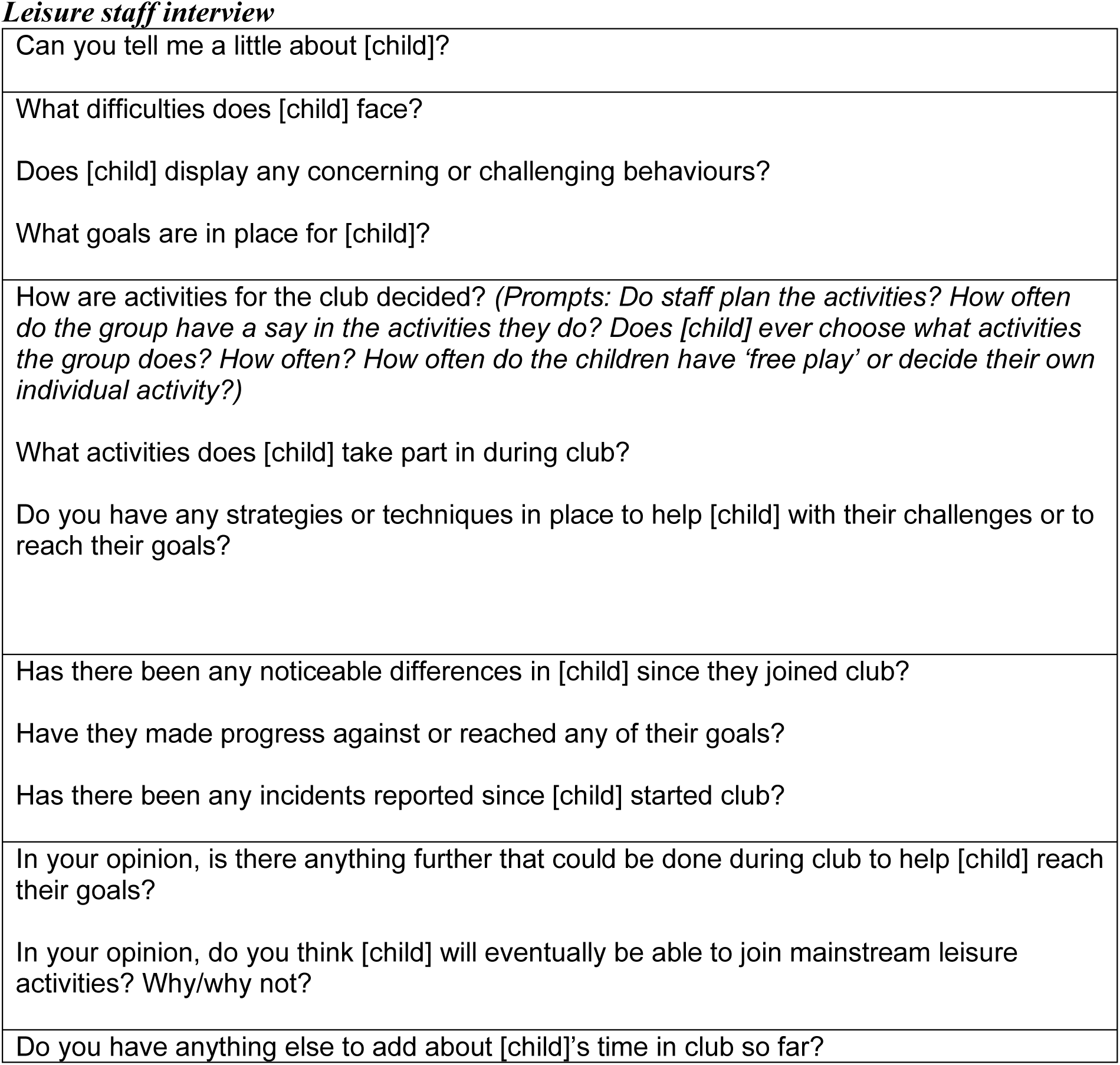
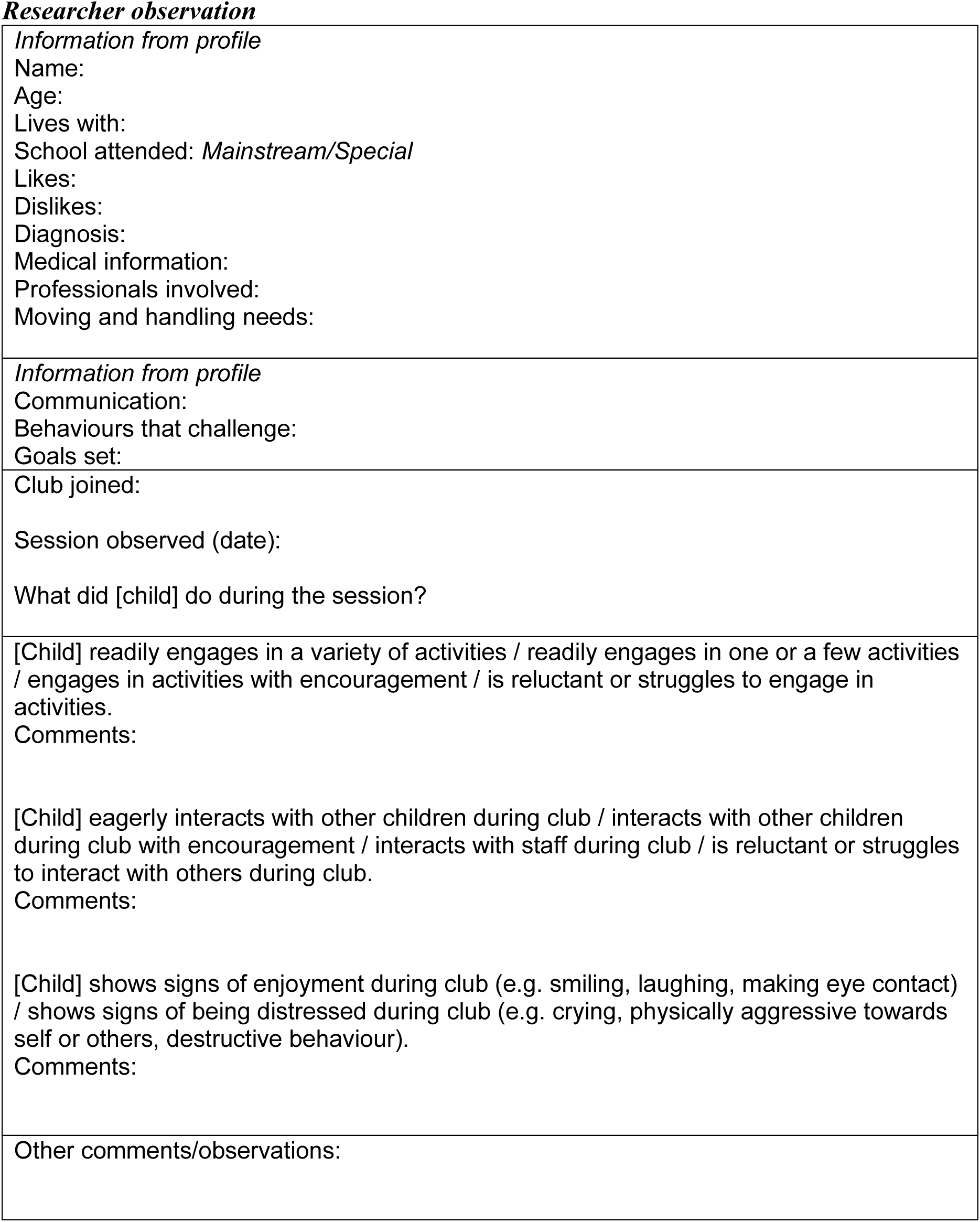

